# Clinical genome sequencing: three years’ experience at a tertiary children’s hospital

**DOI:** 10.1101/2023.02.10.23285786

**Authors:** Runjun D. Kumar, Lisa F. Saba, Haley Streff, Dolores Lopez-Terrada, Jennifer Scull

## Abstract

**Introduction:** Genome sequencing (GS) may shorten the diagnostic odyssey for patients, but clinical experience with this assay in non-research settings remains limited. Texas Children’s Hospital began offering GS as a clinical test to admitted patients in 2020, providing an opportunity to study how GS is utilized, potential for misorders, and outcomes of testing.

**Methods:** We conducted a retrospective review of GS orders for admitted patients for a nearly 3-year period from March 2020 through December 2022. We gathered anonymized clinical data from the electronic health record to answer the study questions.

**Results:** The diagnostic yield over 97 admitted patients was 35%. The majority of GS clinical indications were neurologic or metabolic (61%) and most patients were in intensive care (58%). Tests were often characterized as misordered (56%), frequently due to redundancy with prior testing. Patients receiving GS without prior exome sequencing (ES) had higher diagnostic rates (45%) than the cohort as a whole. In two cases, GS revealed a molecular diagnosis that is unlikely to be detected by ES.

**Conclusions:** The performance of GS in clinical settings likely justifies its use as a first-line diagnostic test, but the incremental benefit for patients with prior ES may be limited.

## Introduction

Clinical genome sequencing (GS) can identify protein-coding single nucleotide variants, as well as intronic, noncoding, regulatory, copy number variants; and repeat expansions through continuous coverage of the nuclear genome. GS may also analyze the mitochondrial genome. GS can provide results faster than other testing methods, especially sequential testing strategies, and may shorten the diagnostic odyssey.

Despite its benefits, clinical experience with GS is still limited. While it is plausible that GS should serve as a first line test for critically ill neonates^1–5^, more general pediatric populations^6–10^, and some adult populations^11,12^, little research addresses the incremental benefit of GS for patients with prior exome sequencing (ES) or chromosomal microarray (CMA)^13^, and current guidelines do not address combining these testing strategies^14^. The diagnostic yield of GS in unselected patient populations is also underexplored^15^.

In 2020, Texas Children’s Hospital (TCH) began offering GS as a clinical test to admitted patients. In 2021 TCH established the Genomic Testing Stewardship Committee (GTSC) to develop evidence-based guidelines for the internal use of GS, ES, CMA, and gene panel testing^16–18^. The GTSC conducted a retrospective review of GS usage at TCH to determine the circumstances in which GS is ordered, the types of misorders that can occur, the diagnostic yield when offered as a clinically reportable test, and how the choice to use GS should be informed by prior testing.

## Materials and Methods

### Sample Identification

We conducted a retrospective review of all genome sequencing (GS) tests performed on patients admitted at TCH from March 2020 through the end of December 2022. Tests were completed on a clinical basis. The review included cancelled tests but excluded tests with pending results. We extracted data from the electronic health record (EHR). The Baylor College of Medicine Institutional Review Board granted a waiver of consent for this study (H-52290).

### Genome Sequencing

We included multiple types of GS in the review, including proband-only, duo (patient + 1 parent), and trio (patient + 2 parents), with both rapid turn-around (7 days) and standard turn-around times (8-10 weeks). All tests were performed at the same CAP/CLIA certified reference laboratory. Molecular findings, diagnoses, and interpretations were obtained exclusively from the final GS report. Tests performed prior to July 22, 2021 did not include triplet nucleotide repeat analysis or mitochondrial genome sequencing; tests after that date include these analyses.

### Data Collection

The GTSC included two laboratory genetic counselors, one clinical molecular geneticist, and one molecular pathologist. Data was collected from the EHR, including GS reports. Major indications for testing were determined from HPO terms in GS reports. Misorders were identified based on clinic notes. The misorder labels included redundant testing, in which prior testing and clinical workup was sufficiently extensive that the GTSC felt more focused testing would have similar utility to GS; controversial testing, in which there is inadequate evidence supporting the use of GS for a given indication; and incorrect order or documentation, where the plan for GS was not clinically documented^16^.

### Statistics

R x64 4.0.3 with base package was used for statistical analyses. P-values represent the outcomes of Fisher exact tests, unless otherwise specified.

## Results

### Circumstances in which GS is ordered

Ninety-seven GS tests for 45 female and 52 male patients were ordered during the study period. The median age was 1.96 years, with a range of 2 days to 25 years old (Figure 1A). Patients were most often on the non-ICU acute care floors (42%), followed by the pediatric ICU (37%), neonatal ICU (12%), and cardiac ICU (8%). The genetics consult service ordered the majority of tests (93%), with neurology being the only other service ordering multiple tests (4%).

**Figure 1.**
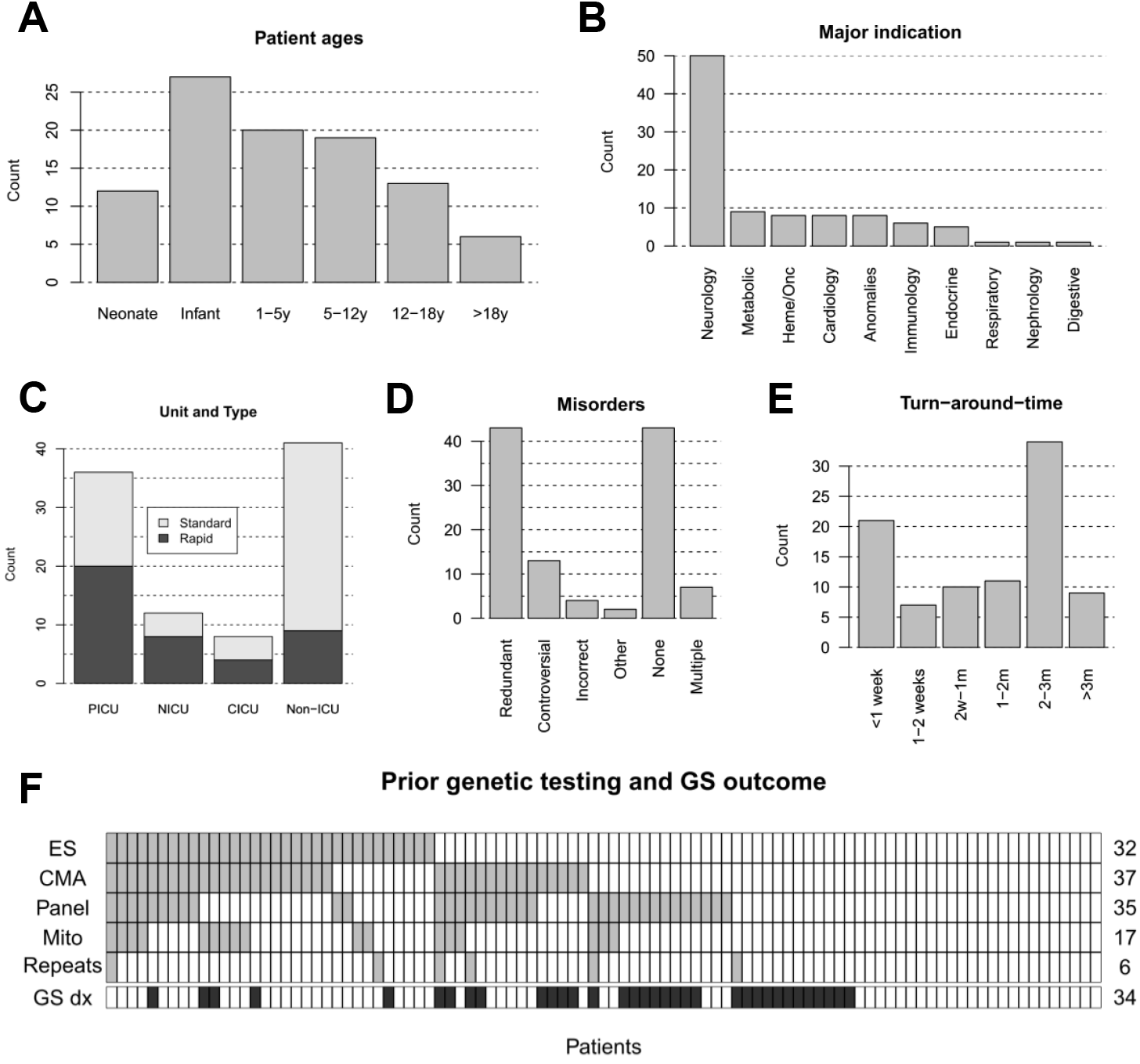
A) Patient ages, n=97. B) Major indication, based on consensus of the GTSC, n=97. C) Admission unit at time of GS order (Pediatric ICU, Neonatal ICU, Cardiac ICU, or non-ICU floors). Rapid and standard turn-around-time orders are indicated, n=97. D) Types of misorders identified; total > 97 due to cases with multiple errors. E) Turn-around times for assays, from time of order to results reported, n=97. F) Prior testing with exome (ES), chromosomal microarray (CMA), targeted DNA panels (Panel), mitochondrial genome sequencing (Mito), and repeat expansion testing (Repeats) are indicated, as well as row totals. Patients with diagnostic genome sequencing (GS dx) are marked. Columns represent patients, n=97.

The most common major indication was neurologic (52%, Figure 1B). Other common indications included metabolic (9.3%), hematologic/oncologic (8.2%), cardiac (8.2%), and multiple congenital anomalies (8.2%). Seizures were the most common subset of neurologic phenotypes (n=36/50), while developmental delays or regression were most common in the remainder (n=9/14). Patients with neurologic phenotypes had a range of ages (2 days to 25 years, median 2.2 years), and were admitted on all unit types (46% acute care, 40% pediatric ICU, 10% neonatal ICU).

### Test appropriateness

Trio GS was completed in 79 cases (81%). Duo GS was completed in 15 cases (15%), and proband-only GS was completed in three cases (4%). Rapid testing, with a laboratory promoted turn-around-time of seven calendar days, was ordered in 41 cases (42%). There is little guidance on when to order rapid testing, and the use of rapid testing depends on clinical context. However, only 9/41 GS orders from acute care floors were requested as rapid, compared with 32/56 from ICUs (22% versus 57%, odds-ratio 4.66 [95% confidence interval 1.8-13.3], p=0.0008), which is consistent with appropriate use in critically ill patients for whom rapid results may guide clinical management.

The GTSC also identified misorders (Figure 1D). Nearly half of orders were found to be appropriate (n=43/97, 44%). The most common concern was testing redundancy (n=43/97, 44%). For these patients, the most common prior tests were ES (n=30/43), CMA (n=29/43), or both ES and CMA (n=21/43). The GTSC suggested these patients could be tested instead with a combination of ES (or ES reanalysis if applicable), CMA, mitochondrial genome sequencing, and repeat expansion testing, depending on the differential diagnosis and prior testing performed.

### Outcomes of testing

GS was cancelled in five cases. For cases 68, 77, and 81, trio GS tests were cancelled per reference lab policy for missing parental samples. For cases 71 and 89, the parents requested test cancellation after the family elected transition to palliative care. For the 92 cases with completed testing, turnaround times were within goal timeframes, accounting for delays in sample collection. Turnaround times show a bimodal distribution, reflecting the combination of rapid and standard orders (Figure 1E).

A diagnosis was achieved in 34 out of 97 tests (35% diagnostic yield, or 37% of completed tests). Fourteen additional tests were “indeterminate”, usually due to a variant of unknown significance (VUS) in a gene linked to the reported phenotype or carrier status in a related gene. As of the time of data collection, no indeterminate cases had been interpreted as diagnostic by the ordering physician. Patients 20, 50, 65, 79, and 93 had prior genetic diagnoses, and GS was ordered to identify potential second diagnoses. GS recapitulated the primary diagnosis in all cases, but only patient 50 received a second diagnosis.

Diagnostic outcome did not vary by unit of admission (p=0.44); the highest diagnostic rate was in the PICU (14/36, 39%), and the lowest in the acute care floors (13/41, 32%). GS had a higher diagnostic rate in patients without prior ES testing compared to those receiving GS after ES (Figure 1F, 29/65 versus 5/32, OR=4.3 [95% CI 1.4-16.1], p=0.006). There was no anti-correlation between GS diagnostic rate and prior CMA, gene panel, mitochondrial genome sequencing, or repeat expansion testing.

### Benefits of genome sequencing over alternative testing

A major goal of the review was to identify scenarios when GS should be recommended after, or instead of, ES. We therefore scrutinized the five patients with positive GS after ES.

For patients 2, 20 and 92, the key molecular finding was previously detected on ES. Patient 2 has *AMMECR1*-associated midface hypoplasia, hearing impairment, elliptocytosis and nephrocalcinosis (MIM# 300990); the P/LP variant was identified on prior ES from 2019 as a VUS. For patient 20, the prior ES actually diagnosed *DIS3L2*-associated Perlman syndrome (MIM# 267000); GS was requested due to suspicion for a second diagnosis. For patient 92, trio GS identified a *de novo* variant in *RYR2* labelled P/LP for *RYR2*-related disease (MIM# 180902). The variant was identified on prior proband-only ES, but was labeled as a VUS due to unknown inheritance.

Patents 87 and 95 had diagnostic GS and prior ES did not report the key molecular result. Patient 87 was diagnosed with Factor XI deficiency (MIM# 612416) based on a P/LP change, c.166T>C, p.C56R in the *F11* gene. ES was completed less than one year prior, but performed at a different reference laboratory. Patient 95 was diagnosed with *TTN*-related disease (MIM# 188840) based on an inherited P/LP change, c.92883G>A, p.W30961*. ES was performed in 2015 and reanalyzed in 2019.

We next sought diagnoses not easily detected by ES or CMA, regardless of prior testing. Patient 18 was diagnosed with *MECOM-*related thrombocytopenia (MIM# 616738) due to a 4kb deletion spanning exon 4 of the gene. Patient 58 was diagnosed with *MT-ND6*-associated MELAS (MIM# 516006) due to a c.221C>T, p.A74V change at 39% heteroplasmy. No patients were diagnosed with structural rearrangements or triplet repeat expansions.

## Discussion

In this retrospective review we characterized the performance of GS when widely available for inpatients admitted in a tertiary children’s hospital. Specifically, we described the circumstances in which GS was ordered, the appropriateness of GS orders, the diagnostic rates of GS in a pediatric hospitalized population, and the benefits of GS over other testing methods.

Prior studies have supported GS use in ICUs, especially NICUs^3,4^. In contrast, the median age of patients receiving GS in this cohort was 2 years old. Moreover, patients were most frequently on the acute care floors and pediatric ICU. Literature generally supports the use of GS for neurologic phenotypes^13^, and the preponderance of neurologic phenotypes in the dataset was expected. Overall, GS testing was used on older, less critically ill children than expected.

We next assessed the appropriateness of GS in individual cases. We found that rapid testing and trio testing were used as expected. However, over half of orders were characterized as misorders, most often due to redundancy with prior testing, especially ES and CMA. Use of GS for patients with extensive prior testing is warranted if GS offers marked incremental benefits, however our analysis does not demonstrate significant incremental diagnostic yield.

The diagnostic rate for the cohort as a whole was 35% (37% of non-cancelled cases). This aligns with prior literature^14^. Use of GS prior to ES was associated with improved diagnostic yield. Only five patients had positive GS after ES, and in three the prior ES detected the P/LP variant. In the remaining two, the identified P/LP variants were coding variants in well-known disease-causing genes which are detectable by ES in principle.

Only two patients from the cohort had GS molecular diagnoses not detectable by ES. One had a single exon deletion in the gene *MECOM*, and the other had a mitochondrial genome variant. Prior studies have argued that GS will improve diagnostic yields by identifying structural variants that are not detectable by ES^13^, but in this cohort no patients fit that description.

Our overall conclusion is that GS has the potential to become a first line genetic test, but its role for patients with extensive prior testing is less clear. Within TCH, the GTSC now recommends that clinicians use GS when three of the following are needed: testing for single gene disorders; testing for copy-number variants; testing for mitochondrial genome disorders; testing for repeat expansion disorders; rapid results. When prior ES is available, reanalysis of exome data may be used prior to GS^19^.

There are important limitations to this study. Foremost, this data is derived from a single center and may not represent the experiences of other facilities. For example, the use of inpatient GS for chronically ill patients may be due to difficulty receiving GS on an outpatient basis, which may not apply to other facilities. Additionally, the diagnostic yield reported here may be improved through the use of RNA-sequencing or functional studies to characterize VUSs, especially in indeterminate cases, and these techniques may be more accessible in other environments^13,20^

Despite these limitations, this study describes the performance of GS in a real-world setting, with clinicians able to order testing without restrictions. The experience of TCH during its first years offering inpatient GS testing may be instructive to other facilities. Additional ongoing test utilization research is needed regarding the diagnostic utility of GS, especially in the context of existing testing paradigms.

## Data Availability

All data described in this study are provided within the article and supplementary materials. Additional de-identified clinical data is available upon request to the corresponding author.

## Author Contribution

Conceptualization: R.D.K., J.S.; Data curation: R.D.K., L.F.S., H.S.; Formal analysis: R.D.K.; Writing – original draft: R.D.K.; Writing – review & editing: R.D.K., J.S., D.L.T., H.S., L.F.S.

## Ethics Declarations

This study adheres to the principles set out in the Declaration of Helsinki. The study was reviewed by the Baylor College of Medicine Institutional Review Board.

## Conflicts of Interest

The authors have no personal conflicts of interest to declare.

---

**Table 1.** The 34 diagnosed patients. Demographic data is shown, as well as high impact molecular findings, including pathogenic (P) and likely pathogenic (LP) variants. Corresponding clinical diagnoses are shown as well as misorder codes (see Methods). Patients 20, 50, 65, 79, and 93 had at least one genetic diagnosis prior to GS.

| Case ID | Age Group | Sex | Test type | TAT (w) | Unit | Major indication | Prior exome | High impact molecular finding | GS diagnosis | Misorder code |
| --- | --- | --- | --- | --- | --- | --- | --- | --- | --- | --- |
| 2 | 1-5y | M | tWGS | 15.6 | PICU | Endocrine | Y | (LP): AMMECR1 c.887+2T>A; (AOH): Chr12 15.6 Mb | Midface hypoplasia, hearing impairment, elipocytosis, and nephrocalcinosis | Redundant |
| 17 | Infant | F | rtWGS | 0.9 | PICU | Neurology | N | (LP): UGDH c.811+1G>A and (VUS): UGDH c.1051A>G, p.I351V | Developmental and epileptic encephalopathy 84 | Redundant |
| 18 | Infant | F | rtWGS | 5.9 | Non-ICU | Heme/Onc | N | (LP): MECOM 4kb exon 4 deletion | Radioulnar synostosis with amegakaryocytic thrombocytopenia 2 | Redundant |
| 20 | 1-5y | M | tWGS | 10.1 | PICU | Anomalies | Y | Homozygous (P): DIS3L2del; (AOH): Chr2 >5Mb | Perlman syndrome | Redundant |
| 26 | Neonate | F | rtWGS | 1.1 | NICU | Metabolic | N | (P): GLDC c.723G>T, p.E575* and (P): GLDC c.2316-1G>A; and secondary finding (P): PMS2 c.825A>G, p.Q275= | Glycine Encephalopathy and Lynch syndrome | NA |
| 28 | Neonate | F | rtWGS | 2.3 | NICU | Anomalies | N | (LP): RAPS N c.133G>A, p.V45M and (LP): RAPS N c.425C>A, p.A142D, (AOH) chrX >5Mb, and secondary finding (P): DSP c.3195C>G, p.Y1065* | Fetal Akinesia deformation sequence 2; Arrhythmogenic Right Ventricular Dysplasia 8 | NA |
| 30 | Neonate | M | rtWGS | 2.7 | NICU | Anomalies | N | (P): CLPB 880C>T p.Q924* and (LP): CLPB c.455+1G>A | 3-methylglutaconic aciduria, type VIIb | Redundant |
| 32 | Infant | F | rtWGS | 4.4 | PICU | Neurology | N | (P): POLR3A c.1771-7C>G and (P): POLR3A c.3781G>A, p.E1261K | Wiedemann Rautenstrauch syndrome | Redundant |
| 33 | 1-5y | M | rtWGS | 0.7 | Non-ICU | Neurology | N | (LP): ARSA c.979+1G>A and (LP): ARSA c.338T>C, p.L113_ | Metochromatic Leukodystrophy | NA |
| 34 | 5-12y | F | tWGS | 23.7 | Non-ICU | Immunology | N | (P): NKFBIA c.95_96delinsTT, p.S32I | Ectodermal dysplasia and immunodeficiency 2 | NA |
| 35 | Neonate | F | rdWGS | 1.1 | CICU | Neurology | N | (P): KCNH2 c.991del, p.S331Afs*29 | Long QT syndrome 2 | NA |
| 36 | 1-5y | M | tWGS | 10 | Non-ICU | Neurology | N | (P): PNPT1 c.1519G>T, p.A507S and (LP): PNPT1 c.1592C>G, p.T531R | Combined oxidative phosphorylation deficiency 13 | Redundant |
| 37 | 5-12y | F | dWGS | 12 | Non-ICU | Cardiology | N | (P): PKP2 c.1926T>A, p.Y642* | Arrhythmogenic right ventricular dysplasia 9 | Controversial |
| 41 | Infant | F | rtWGS | 1.7 | PICU | Heme/Onc | N | (P): RPS26 c.9_12del, p.K4Efs*40 | Diamond Blackfan Anemia 10 | NA |
| 45 | 12-18y | F | tWGS | 9 | Non-ICU | Neurology | N | (P): ATP1A3 c.2305C>T, p.R769C | Dystonia-12 | NA |
| 48 | Infant | F | rtWGS | 0.7 | PICU | Immunology | N | (P): FLCN c.250-2A>G | Birt-Hogg Dube | Controversial |
| 50 | 12-18y | F | rdWGS | 1.4 | PICU | Neurology | N | (P): chr22del and (LP): TYMP c.1040T>C, p.L347P | Phelan McDermid and Mitochondrial DNA Depletion Syndrome 1 (MNGIE) | Redundant |
| 52 | Infant | M | rtWGS | 0.6 | PICU | Immunology | N | (P): NDUFS7 c.364G>A, p.V122M and (LP): NDUFS7 c.537C>A, p.Y179* | Mitochondrial complex I deficiency, nuclear type 3 (Leigh syndrome) | NA |
| 54 | Neonate | M | rdWGS | 2.3 | NICU | Neurology | N | (P): ARX c.769del, p.R257Afs*68 | Proud syndrome | Controversial |
| 56 | 1-5y | M | rtWGS | 0.9 | PICU | Neurology | N | (LP): OFD1 c.991C>T, p.Q331* | Joubert syndrome, 10 | NA |
| 58 | 12-18y | F | tWGS | 10 | Non-ICU | Endocrine | N | (P): MT-ND6 c.221C>T, p.A74V (39% heteroplasmy); AOH >5Mb | MT-ND6 related disorders (MELAS) | NA |
| 59 | 1-5y | F | dWGS | 9.9 | Non-ICU | Neurology | N | (P): ANKRD11 c.928_934del, p.P310Vfs*56 | KBG syndrome | Multiple |
| 65 | 1-5y | F | tWGS | 10 | CICU | Cardiology | N | (P): 7q11.23del | Williams syndrome | NA |
| 69 | 1-5y | M | rtWGS | 0.6 | PICU | Neurology | N | (P): RARS2 c.419T>G, p.F140C and (LP): RARS2 c.36+1G>T | Pontocerebellar hypoplasia, RARS2 related | NA |
| 76 | Infant | M | rtWGS | 3.1 | PICU | Immunology | N | (P): F8 c.1569 G>T, p.L523= | Hemophilia A | NA |
| 79 | 12-18y | F | dWGS | 9.9 | Non-ICU | Anomalies | N | (P): OCA2 c.1211C>T, p.T404M and (P): OCA2 c.1327G>A, p.V443I | Oculocutaneous albinism II | NA |
| 83 | Infant | M | tWGS | 10.3 | PICU | Endocrine | N | (P): NF1 c.4812C>A, p.Y1604* | Neurofibromatosis type 1 | Controversial |
| 84 | Infant | F | rtWGS | 0.4 | PICU | Neurology | N | (P): ATP1A2 c.1022G>A, p.C341Y | ATP1A2-related disorders | NA |
| 87 | Infant | M | tWGS | 10 | Non-ICU | Heme/Onc | Y | (LP): F11 c.166T>C, p.C56R | Factor XI deficiency | Redundant |
| 88 | 12-18y | M | pWGS | 14.6 | CICU | Neurology | N | (P): DMD c.6439-636_7098+908del (exon 45-48 del) | Duchenne/Becker muscular dystrophy | NA |
| 92 | 12-18y | F | tWGS | 10 | Non-ICU | Neurology | Y | (LP): RYR2 c.11805G>C, p.L3935F | RYR2-Related disorders | Redundant |
| 93 | 12-18y | F | tWGS | 7.3 | Non-ICU | Anomalies | N | (P): dup(21)(p21q22) | Down syndrome | NA |
| 94 | 5-12y | F | tWGS | 10.9 | PICU | Neurology | N | (LP): MYBPC3 c.3407_3409del, p.Y1136del | MYBPC3-related disorders | Multiple |
| 95 | 5-12y | M | tWGS | 7 | Non-ICU | Metabolic | Y | (LP): TTN c.92883G>A, p.W30961* | TTN-related disorders | Redundant |

## Notes

### Competing Interest Statement

The authors have declared no competing interest.

### Funding Statement

This study did not receive any funding.

### Author Declarations

The Baylor College of Medicine Institutional Review Board granted a waiver of consent for this study (H-52290).

